# One-Year Brain Structural Changes Are Associated with Postoperative Delirium and Delayed Resolution of Interleukin-6

**DOI:** 10.64898/2026.05.03.26352074

**Authors:** Jinglei Lv, Jennifer Taylor, Samantha Curtis, Kaitlin Kramer, David Kunkel, Shalini Thakur, Veena Nair, Matthew I. Banks, Robert A. Pearce, Vivek Prabhakaran, Richard Lennertz, Robert D. Sanders

## Abstract

**Background:** Postoperative delirium is a common complication in older adults and is associated with neuroinflammation and cognitive decline. Previous studies have shown that the number of surgical procedures is associated with hippocampal volume loss in older adults in a large-scale UK Biobank study. However, it remains unclear whether hippocampal volume loss within one year after surgery is associated with postoperative delirium.

**Methods:** Longitudinal structural MRI data and blood biomarkers were collected before surgery and one year postoperatively from 62 participants (>65 years, 27 females) undergoing major non-intracranial surgery. Hippocampal and other subcortical volumes were quantified using FreeSurfer. Cortical thickness was measured for cortical regions defined by the Desikan–Killiany (DK) atlas. One-year structural changes were examined in relation to peak Delirium Rating Scale (DRS) scores and one-year changes in plasma interleukin (IL)-6 levels.

**Results:** One-year volume loss in the right hippocampus was significantly correlated with postoperative peak DRS scores and the one-year change in IL-6. Additional gray matter reductions were observed in the right putamen and the right superior parietal cortex. Right putamen volume loss was also associated with the one-year change in IL-6, while cortical thinning in the right superior parietal cortex was associated with peak DRS scores.

**Conclusions:** Postoperative delirium is associated with longitudinal gray matter loss following surgery. Delayed resolution of inflammation may also contribute to postoperative brain structural changes.

**Clinical trial registration:** NCT01980511 and NCT03124303.

## 1. Introduction

Postoperative delirium is a common and serious complication in older surgical patients, characterised by acute disturbances in attention, cognition, and arousal. It affects a substantial proportion of older adults undergoing major surgery and is associated with increased mortality, prolonged hospitalisation, and long-term cognitive decline[1,2]. Population-scale neuroimaging studies have suggested that surgical exposure may be associated with accelerated (average) hippocampal atrophy in older adults [3]. Consistent with this, evidence suggests that surgery and anaesthesia may be associated with markers of neuronal injury[4] and subsequent cognitive decline[3,5,6]. Accumulating data suggest that delirium represents a manifestation of perioperative neuronal injury[4,7] rather than merely a transient behavioural disturbance. Studies of neurofilament light have demonstrated perioperative neuronal injury and have been linked to delirium severity[8]. However, the structural neuroanatomical consequences of postoperative delirium remain incompletely characterised. Given the associations of surgical exposure with hippocampal degeneration, we hypothesized that atrophy would be proportional to the severity of delirium postoperatively.

Neuroinflammation is widely considered a central mechanism underlying postoperative delirium[9]. Surgical trauma triggers a systemic inflammatory response that can disrupt the blood–brain barrier and activate neuroimmune pathways within the central nervous system. This process may allow peripheral inflammatory mediators to access the brain and trigger microglial activation and neuronal injury. Clinical studies have shown that biomarkers reflecting blood–brain barrier dysfunction and neuroinflammatory responses are associated with the incidence and severity of postoperative delirium[10,11]. Pro-inflammatory cytokines, including interleukin-6 (IL-6), are frequently elevated following surgery and have been implicated in delirium pathogenesis through their effects on neurovascular integrity and synaptic function. We recently showed in this cohort that delayed resolution of inflammation following surgery was associated with delayed cognitive recovery[12], however, it is unknown if there was a structural imprint on the brain of this prolonged inflammatory response. The hippocampus is of particular interest in this context because of its critical role in memory and vulnerability to inflammatory and metabolic insults. However, whether hippocampal volume loss, following surgery, is associated with delirium severity or inflammatory recovery remains unclear.

In this study, we investigated longitudinal brain structural changes in older adults undergoing major non-intracranial surgery. Using pre- and one-year postoperative structural MRI, we examined whether postoperative delirium severity and recovery of systemic inflammation, measured by IL-6, were associated with changes in hippocampal volume and other brain structures. We hypothesised that greater delirium severity and persistent inflammatory responses would be associated with greater longitudinal average hippocampal volume loss. Exploratory analyses then investigated equal affects in right or left hippocampus and other regions with gray matter loss.

## 2. Method

### 2.1 Data collection

Data were obtained from the prospective study Interventions for Postoperative Delirium: Biomarker-3 (IPOD-B3, ClinicalTrials.gov: NCT03124303), approved by the Institutional Review Board of the University of Wisconsin–Madison (2015-0374). MRI scans and blood samples for biomarker analyses were collected both before and one year after surgery. Surgical procedures included cardiovascular, spinal, general, and urological operations. Individuals were excluded if they were unable to communicate with research staff because of language or sensory barriers, had a documented history of dementia, or resided in a skilled nursing facility. All participants provided written informed consent before participation.

### 2.2 Delirium Assessment

Delirium symptom severity was evaluated twice daily on postoperative days 1–4 using the Delirium Rating Scale–98 (DRS-98), which provides a quantitative measure of delirium symptom severity. Delirium status (present/absent) was assessed by the 3D-CAM. For intubated patients, delirium was evaluated using the CAM-ICU in combination with the Richmond Agitation–Sedation Scale (RASS). Patients who had a CAM-ICU performed were scored for delirium severity after extubation on the DRS-98.

### 2.3 Imaging

Structural MRI data were acquired on three 3 T General Electric MRI scanners, i.e., scanner 1: 3T GE750, Scanner2: 3 T Signa PETMRI, and scanner 3: 3T GE750, using an eight-channel head coil. High-resolution T1-weighted images were collected with a FSPGR BRAVO sequence (TR = 8.132 ms, TE = 3.18 ms, TI = 450 ms, 256 × 256 matrix, 156 slices, flip angle = 12°, FOV = 25.6 cm, slice thickness = 1 mm). Structural images were processed using FreeSurfer (version 8.1). The standard recon-all pipeline was applied, which includes skull stripping, spatial registration, tissue segmentation, and cortical parcellation. Cortical thickness measurements were derived using the Desikan–Killiany (DK) atlas, comprising 68 cortical regions (34 per hemisphere). Subcortical volumes were obtained from the automated segmentation generated by the FreeSurfer processing pipeline. The volume was normalized as the percentage of intracranial volume.

### 2.4 Blood Biomarker

Blood samples were drawn into EDTA-containing tubes and stored at −80 °C until analysis. Plasma levels of interleukin-6 (IL-6) were quantified using Nucleic acid Linked-Sandwich Assay (NULISA) conducted at university of Wisconsin. Samples were collected at baseline, on postoperative days (PODs) 1–9, and at 90 days and one year following surgery.

### 2.5 Statistics

Statistics were performed using MATLAB. Pearson’s correlation was first used to assess the relationships between one-year hippocampal volume change (Δ = 1 year − baseline) and both peak Delirium Rating Scale (DRS) scores and one-year changes in IL-6. Our primary outcome was the effects of delirium severity and IL-6 on average hippocampal volume. Secondary outcomes included associations with left and right hippocampus and. Tertiary analyses focussed on subcortical volumes and cortical thickness that changed over the first postoperative year.

Linear mixed-effects models were fitted as follows:

Hippo volume ∼ 1 + Time +Peak DRS + Age + Gender + Scanner+ Time: Peak DRS

+ (1 | SubjectID)

Hippo volume ∼ 1 + Time + IL6 + Age + Gender + Scanner+ Time: IL6 + (1 | SubjectID)

In these models, hippocampal volume was treated as the dependent variable, with peak DRS or IL-6 as fixed effects, and subject-specific intercepts included as random effects. Interaction terms between time and the variable of interest were included to capture longitudinal effects. Separate models were performed for the bilateral average hippocampal volume, as well as for the left and right hippocampus individually. Preoperative cognitive variables were not included in the model as they would be at least partly dependent on hippocampal volume.

Group-level paired t-tests were conducted to assess one-year changes in cortical thickness and subcortical volumes. Multiple comparisons were controlled using the false discovery rate (FDR) with a significance threshold of p < 0.05. For regions showing significant structural changes, Pearson’s correlation was further used to examine associations with peak DRS scores and one-year changes in IL-6.

## 3. Results

A total of 296 patients aged ≥65 years undergoing elective major surgery with an expected hospital stay of ≥2 days were recruited. Among them, 142 participants had preoperative MRI scans, and 282 of them had blood samples. 70 participants underwent MRI scans 1 year post operation, and 150 of them had blood test. In this study, we only included the 62 participants (27 females) who have both longitudinal MRI scans and blood tests (Fig.1).

**Figure 1.**
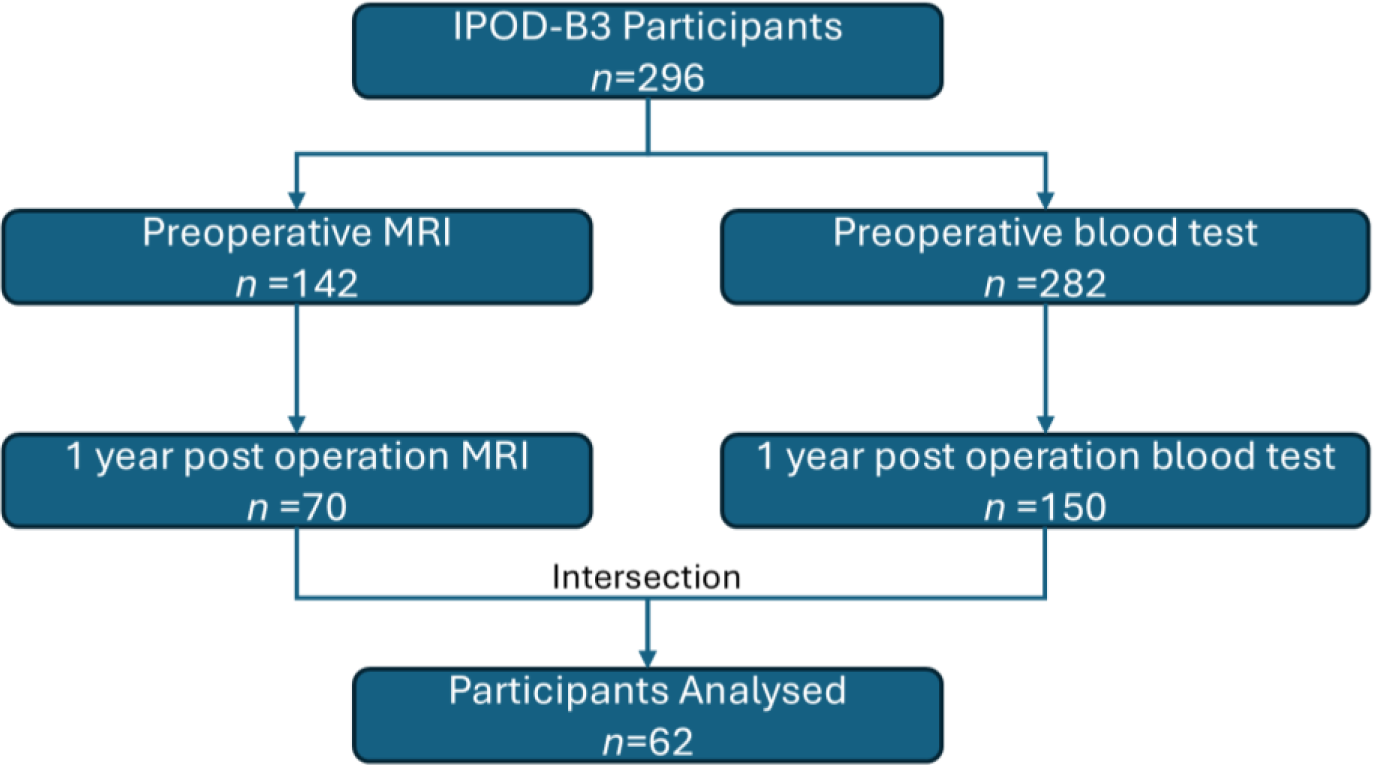
STROBE plot depicts the relevant participants from recruitment to analysis. Only subjects with both longitudinal MRI scans and blood tests were included.

### 3.1 Hippocampal Volume Loss Is Associated with Peak Delirium Severity and Delayed Resolution of IL6

Given prior evidence that the hippocampus is particularly vulnerable to surgical exposure[3], we first examined whether hippocampal volume change over one year was associated with postoperative delirium severity. We also hypothesized that persistent neuroinflammation may contribute to long-term hippocampal atrophy.

As shown in Fig. 2 and Tables 1–2, mixed-effects model analyses demonstrated that greater one-year hippocampal volume loss was significantly associated with peak DRS scores. This result is particularly dominated by right hippocampus (p<0.05, Fig.2a-b), while not significant with left hippocampus (Fig.2c). In addition, raised IL-6 at one year relative to the preoperative baseline was significantly associated with hippocampal volume loss, for both left and right side (p<0.05, Fig.2d-f), though mixed effects modelling suggested that interaction effect is only significant on the right.

**Figure 2.**
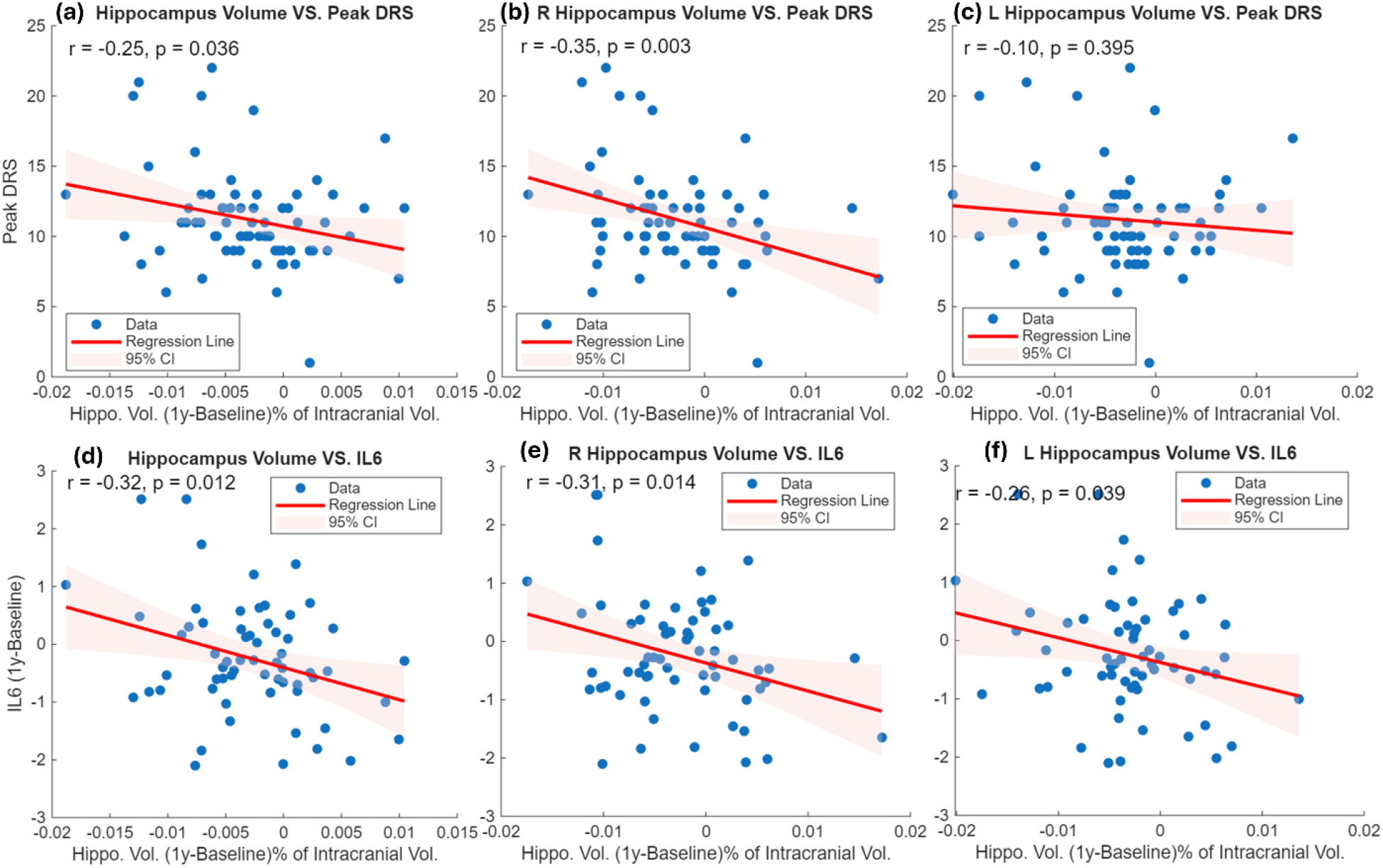
Association between one-year hippocampal volume change (1 year − baseline) and postoperative delirium severity and one year change of IL-6 (log2 transformed). (a–c) Relationships between peak Delirium Rating Scale (DRS) scores and volume change in the bilateral hippocampus (average), right hippocampus, and left hippocampus. (d–f) Relationships between one-year change in IL-6 (1 year − baseline) and volume change in the bilateral hippocampus (average), right hippocampus, and left hippocampus.

**Table 1.** Linear mixed effects model for hippocampus volume and peak DRS.

| Region | Variable | $\beta$ (95% CIs) | t | DF | p-value |
| --- | --- | --- | --- | --- | --- |
| Hippocampus (Average) | Time | 0.00185 (-0.00239, 0.00609) | 0.865 | 116 | 0.389 |
|  | Peak DRS | 0.000714 (-0.000991, 0.00242) | 0.829 | 116 | 0.409 |
|  | Age | -0.00201 (-0.00334, -0.000676) | -2.99 | 116 | 0.003 |
|  | Sex | 0.0173 (0.0044, 0.0302) | 2.658 | 116 | 0.009 |
|  | Scanner1 | -0.0228 (-0.056, 0.0104) | -1.36 | 116 | 0.176 |
|  | Scanner2 | -0.0026 (-0.0198, 0.0146) | -0.3 | 116 | 0.765 |
|  | **Time: Peak DRS** | **-0.00042 (-0.000782, -5.79e-05)** | **-2.3** | **116** | **0.023** |
| Right Hippocampus | Time | 0.00443 (-0.00022, 0.00907) | 1.887 | 116 | 0.062 |
|  | Peak DRS | 0.000273 (-0.00158, 0.00212) | 0.292 | 116 | 0.771 |
|  | Age | -0.00198 (-0.00342, -0.000534) | -2.71 | 116 | 0.008 |
|  | Sex | 0.017 (0.003, 0.0309) | 2.406 | 116 | 0.018 |
|  | Scanner1 | -0.026 (-0.0621, 0.01) | -1.43 | 116 | 0.155 |
|  | Scanner2 | -0.000513 (-0.0192, 0.0182) | -0.05 | 116 | 0.957 |
|  | **Time: Peak DRS** | **-0.00063 (-0.00103, -0.000234)** | **-3.15** | **116** | **0.002** |
| Left Hippocampus | Time | -0.000724 (-0.00548, 0.00403) | -0.3 | 116 | 0.764 |
|  | Peak DRS | 0.00115 (-0.000571, 0.00288) | 1.325 | 116 | 0.188 |
|  | Age | -0.00204 (-0.00338, -0.00069) | -3 | 116 | 0.003 |
|  | Sex | 0.0176 (0.00457, 0.0306) | 2.676 | 116 | 0.009 |
|  | Scanner1 | -0.0196 (-0.0532, 0.014) | -1.16 | 116 | 0.250 |
|  | Scanner2 | -0.00469 (-0.0221, 0.0127) | -0.53 | 116 | 0.595 |
|  | **Time: Peak DRS** | **-0.000209 (-0.000615, 0.000196)** | **-1.02** | **116** | **0.309** |

**Table 2.** Linear mixed effects model for hippocampus volume and IL6 (log2 transformed).

| Region | Variable | $\beta$ (95% CIs) | t | DF | p-value |
| --- | --- | --- | --- | --- | --- |
| Hippocampus (Average) | Time | 0.00197 (-0.0152, 0.0191) | 0.227 | 116 | 0.821 |
|  | **IL6** | **-0.00169 (-0.00321, -0.000173)** | **-2.21** | **116** | **0.029** |
|  | Age | -0.00193 (-0.00325, -0.000605) | -2.89 | 116 | 0.005 |
|  | Sex | 0.017 (0.00425, 0.0298) | 2.638 | 116 | 0.009 |
|  | Scanner1 | -0.0224 (-0.0553, 0.0105) | -1.35 | 116 | 0.179 |
|  | Scanner2 | -0.000919 (-0.0179, 0.0161) | -0.11 | 116 | 0.915 |
|  | Time:IL6 | -0.000413 (-0.00176, 0.00093) | -0.61 | 116 | 0.544 |
| Right Hippocampus | **Time** | **0.0194 (0.000691, 0.038)** | **2.054** | **116** | **0.042** |
|  | IL6 | -0.00123 (-0.00289, 0.000425) | -1.47 | 116 | 0.144 |
|  | Age | -0.00187 (-0.00329, -0.000441) | -2.59 | 116 | 0.011 |
|  | Sex | 0.017 (0.0032, 0.0308) | 2.441 | 116 | 0.016 |
|  | Scanner1 | -0.0268 (-0.0623, 0.00858) | -1.5 | 116 | 0.136 |
|  | Scanner2 | 0.0004 (-0.0179, 0.0187) | 0.043 | 116 | 0.966 |
|  | **Time:IL6** | **-0.00177 (-0.00323, -0.000304)** | **-2.39** | **116** | **0.018** |
| Left Hippocampus | Time | -0.0154 (-0.0342, 0.00329) | -1.63 | 116 | 0.105 |
|  | **IL6** | **-0.00216 (-0.00382, -0.000502)** | **-2.58** | **116** | **0.011** |
|  | Age | -0.00199 (-0.00335, -0.000632) | -2.9 | 116 | 0.004 |
|  | Sex | 0.0171 (0.00398, 0.0302) | 2.581 | 116 | 0.011 |
|  | Scanner1 | -0.018 (-0.0518, 0.0157) | -1.06 | 116 | 0.292 |
|  | Scanner2 | -0.00223 (-0.0197, 0.0152) | -0.25 | 116 | 0.800 |
|  | Time:IL6 | 0.000942 (-0.000526, 0.00241) | 1.271 | 116 | 0.206 |

### 3.2 Volume Loss on other subcortical structures

In exploratory analyses, we used paired t-statistics to examine volume changes in 17 subcortical structures. Significant volume loss (FDR-corrected, p < 0.05) was observed in the left hippocampus, right hippocampus, left putamen, and right putamen (Fig. 3a). However, putamen volume loss was not associated with delirium severity (Fig. 3b–c). In contrast, right putamen volume loss was associated with the persistence of IL-6 (Fig. 3d).

**Figure 3.**
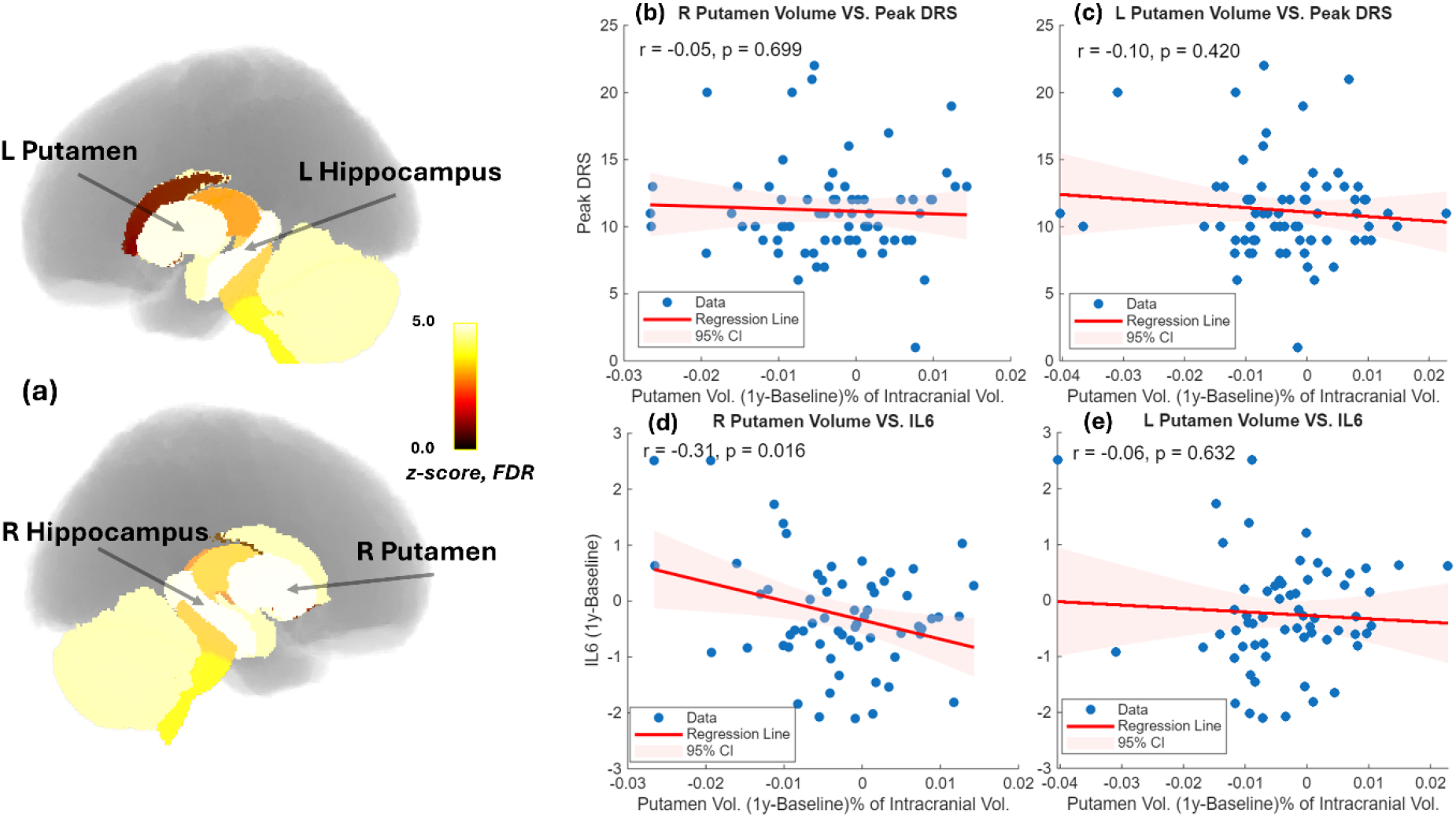
(a) Significant 1 year volume loss was found with left hippocampus, right hippocampus, left putamen, and right putamen. (b) Right putamen volume loss vs peak DRS. (c) Left putamen volume loss vs peak DRS. (c) Right putamen volume loss vs IL6 recovery. (d) Left putamen volume loss vs IL6 recovery.

### 3.3 One-year Cortical Thinning

Next, we extended to study regional cortical thickness. Longitudinal analysis revealed significant one-year cortical thinning in the left caudal middle frontal, left rostral middle frontal, right precentral, and right superior parietal cortices (FDR-corrected, p < 0.05, Fig.4a). Among these regions, cortical thinning in the right superior parietal cortex was significantly associated with peak DRS scores (Fig.4b). However, it is not correlated with the one-year change of IL6 (Fig.4c). No significant correlations were found for the other cortical regions (Fig.4d-i).

**Figure.4.**
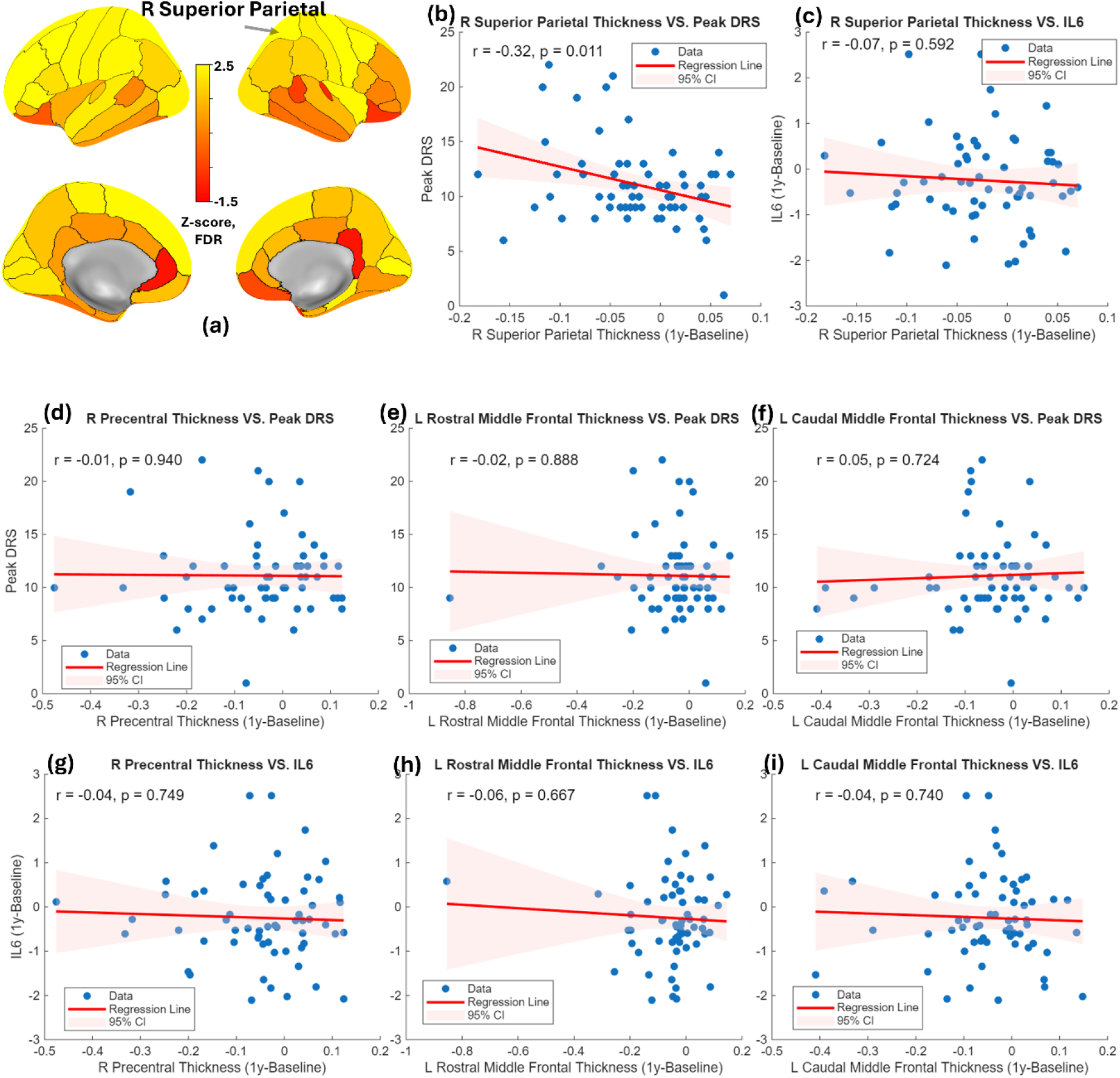
(a) Z-score map illustrating regions of significant one-year cortical thinning based on the Desikan–Killiany (DK) atlas. (b) Association between one-year change in right superior parietal cortical thickness and peak DRS. (c) Association between one-year change in right superior parietal cortical thickness and one-year change in IL-6. (d–f) Associations between one-year cortical thinning in other significant thinning regions and peak DRS scores. (g–i) Associations between one-year cortical thinning in other significant thinning regions and one-year change in IL-6.

## 4. Discussion

In this longitudinal study, we investigated the relationship between postoperative delirium severity, systemic inflammation, and one-year structural brain changes following surgery. Our results show that greater hippocampal volume loss over one year is associated with higher postoperative delirium severity, and delayed resolution of IL-6. These findings suggest that postoperative delirium and persistent inflammatory responses may contribute to long-term structural brain alterations.

The hippocampus plays a critical role in memory and cognitive processing and is particularly vulnerable to metabolic stress and inflammatory injury. Previous studies have suggested that delirium may accelerate cognitive decline and increase the risk of subsequent dementia [13,14]. Our findings extend this evidence by demonstrating that delirium severity is associated with longitudinal hippocampal volume loss, supporting the hypothesis that delirium reflects or contributes to injury within memory-related neural systems.

Notably, the association was particularly evident in the right hippocampus, that may suggest hemispheric differences in vulnerability. The right hippocampus has been implicated in spatial cognition, contextual integration, and attentional processing, domains frequently disrupted during delirium. Previous neuroimaging studies have also reported asymmetric hippocampal involvement in aging and neurodegenerative conditions [15,16]. The preferential association with the right hippocampus observed here may therefore reflect selective disruption of neural circuits supporting attention, spatial orientation and situational awareness, which are core features of delirium. Alternatively, lateralized vulnerability may arise from differential sensitivity of hippocampal subfields to inflammatory and metabolic stress. An alternative explanation is that changes in the right hippocampus (which is larger than the left) are more easily detected over the 1-year time period. We also identified cortical thinning in frontal and parietal regions, including the right superior parietal cortex, which showed a significant association with delirium severity. The superior parietal cortex is involved in attention, particularly spatial, and sensory integration, cognitive domains frequently impaired in delirium. [17] Overall, the changes imply that following surgery cognitive tests should include spatial navigation tasks as this is an important function for people and a key part of completing activities of daily living.

Inflammation has been widely proposed as a key mechanism underlying postoperative cognitive decline. Elevated levels of interleukin-6 (IL-6) have been linked to delirium risk and neuronal injury [18,19]. Consistent with this hypothesis, we found that delayed resolution of IL-6 was associated with hippocampal volume loss, suggesting that persistent inflammatory responses following surgery may contribute to long-term structural brain changes. Experimental studies indicate that inflammatory cytokines can impair synaptic plasticity, suppress neurogenesis, and promote neurodegenerative processes within the hippocampus [20]. It is of interest that delayed resolution of IL-6 did not show a hemispheric specific vulnerability while, delirium severity does. If validated in future studies, this implies separate mechanisms of injury.

Beyond hippocampal changes, we observed significant longitudinal volume loss in the putamen, although this was not associated with delirium severity. Interestingly, right putamen volume loss was associated with delayed resolution of IL-6, indicating that inflammatory processes may influence broader subcortical structures beyond limbic regions. However, this was a tertiary analysis and so should be considered hypothesis generating, not testing.

Several limitations should be considered. First, the observational design limits causal inference regarding the relationships between delirium, inflammation, and structural brain changes. Second, there was significant loss to follow up in our imaging cohort that may introduce selection bias. It is likely that more severely affected individuals may be more likely to be lost to follow up but this is known. Third, we concentrate on IL-6, having previously shown an effect on cognition in our cohort[12] and to avoid false positives from testing multiple cytokines or other biomarkers. Nonetheless, additional biomarkers may provide a more comprehensive characterization of postoperative neuroinflammatory responses. Finally, replication in larger cohorts will be necessary to confirm our findings.

In summary, this study demonstrates that postoperative delirium severity and inflammatory recovery trajectories are associated with long-term structural brain changes, particularly right hippocampal atrophy. These findings support the hypothesis that neuroinflammation may mediate postoperative brain vulnerability and highlight the hippocampus as a key target for understanding the long-term neurological consequences of delirium and surgical recovery.

## Data Availability

All data produced in the present work are contained in the manuscript

